# A Prospective Cohort Study to Develop Multi-Biomarkers Panel to Define Biological Ageing in Five Different Cohorts from Newborn to Oldest Adult: A Study Protocol

**DOI:** 10.1101/2025.03.17.25324084

**Authors:** Prasun Chatterjee, Rashi Jain, Pooja, Avinash Chakrawarty, Lata Rani, Sharmistha Dey, Rashmita Pradhan, Vidushi Kulshrestha, Lakshmy Ramakrishnan, Prasun Chatterjee

## Abstract

**Background:** Age-associated disease management depends significantly on chronological age and macro-level clinical data sets. However, the biological age captures bio-physiological deterioration more precisely than the calendar age. Biological ageing is the accumulation of successive damage to various cells, tissues, and individual organs over the ageing period. Quantifying biological ageing could be of great value for better clinical decision-making. Various epigenetic clocks, including the Hannum clock, GrimAge clock, Horvath clock, PhenoAge clock, and DunedinPACE, have been used to quantify biological age. However, epigenetics alone cannot explain all other critical processes, ranging from ageing hallmarks, signalling pathways, clinical phenotypes, physiological functioning, and environmental exposure to lifestyle habits that participate in the ageing process.

Therefore, our primary objective is to define reliable, reproducible, robust, and integrative biomarkers that can manifest all ageing hallmarks and other associated factors to quantify biological age.

**Methods/Design:** This community-based prospective cohort study will be conducted at the National Centre of Ageing, All India Institute of Medical Sciences, New Delhi. This study will include 200 participants from five cohorts, *i*.*e*. newborns, adolescents (10-19 years), middle-aged individuals (20-59 years), young olds (60-79 years), and the oldest old (Above 80 years). Forty individuals from each cohort will be recruited to study blood and stool biomarkers along with a comprehensive assessment of cognitive behavior, psychological well-being, functional capacity, gut health, nutritional behaviour and physiological measures. Participants will also be monitored in real-time through wearable devices. After five years, participants will be followed up with the same biomarkers to gain insights about the speed of ageing, predicting disease and mortality. Multi-domain data will be integrated to develop a deep learning-based multi-model algorithm for biological age estimation.

**Conclusion:** This first-of-its-kind study would provide an exhaustive understanding of the ageing process throughout life, 0-100 years. Integrative biomarkers would make a precise determination of biological age. Additionally, the study of change in these parameters after five years would elucidate the pace of biological ageing and predict life expectancy and disability.

## Introduction

Ageing is a gradual, incessant, natural process that involves heterogeneous changes in dynamic biological, physiological, psychological, environmental, behavioural, and social processes (1). Chronological or calendar age is generally considered a defining criterion along with macro clinical data sets in clinical decision-making. The correlation between chronological age and physical ageing is readily apparent to clinicians. However, biological ageing, which refers to the ageing of various cells, tissues, and individual organs, is the more precise determinant of health (2). It captures bio-physiological deterioration more precisely than calendar age. Exploring and unravelling the basis of biological ageing is a significant scientific challenge.

Despite the remarkable progress and enhanced scientific interest in this field in the last decade, biomarkers to explain biological age and the ageing process have not been validated yet (3). We need reliable, relevant, reproducible, and robust biomarkers to quantify biological damage accumulation during ageing. As per The American Federation for Aging guidelines (AFAR), a promising ageing biomarker should indicate ageing processes and age-associated outcomes and be responsive to an exposure or intervention, including therapeutic interventions (4).

Blood-based molecular biomarkers are the most extensive and precise among all known clinical, molecular, and physiological biomarkers (5). They comprehensively reflect physiological and pathological states due to their accessibility, high sensitivity, and ability to capture systemic changes across the body. Consequently, Omics approaches, specifically epigenetic identification of age-associated molecules, are considered the primary biomarker of biological ageing (6,7).

Over the past decade, three generations of epigenetic clocks have evolved: first-generation: trained to assess chronological age based on DNA methylation pattern (8,9), second-generation: trained to calculate biological age based on the correlation between methylation and health risks data (10,11), and the third-generation: integrates ageing signs and organ-specific health parameters with methylation data to measure the pace of ageing (12).

However, epigenetic alteration represents only one of the twelve hallmarks of ageing described by Lopez et al. in their seminal paper (13,14).We need biomarkers that can manifest all these hallmarks which are genomic instability (GI), telomere attrition (TA), epigenetic alterations (EA), loss of proteostasis (LP), deregulated nutrient-sensing (DNS), mitochondrial dysfunction (MD), cellular senescence (CS), stem cell exhaustion (SCE), altered intercellular communication (AIC), chronic inflammation (CI), dysbiosis (DB), and disabled macroautophagy (DM).

All these hallmarks are significantly impacted by the plethora of critical factors ranging from clinical phenotypes, physiological functioning, and environmental exposure to lifestyle habits (15). Hence, we planned to explore the ageing biomarkers that can elucidate all twelve hallmarks and their associated factors based on all-inclusive data from blood, stool, physiological assessments, psychological assessments, functional performance, living conducts, and real-time digital monitoring. The secondary aim is the integration of all these biomarkers to create a deep learning-based multi-model algorithm for biological age estimation. These integrative biomarkers condense the signal even from the least expressed parameters to generate robust data and new knowledge.

Therefore, our objective with this first-of-its-kind study is to evaluate the age-sensitive multi-parametric biomarkers in five cohorts from 0 to 100 years consisting of newborns, adolescents, middle-aged, young old and oldest old people to define biological ageing precisely. This would provide an exhaustive understanding of the ageing process throughout life. The change in these parameters will also be studied after 5 years to assess the pace of biological ageing and predict life expectancy and disability.

## Methodology

### Study Design and Population

This community-based prospective cohort study has been designed at the National Centre of Ageing, All India Institute of Medical Sciences, New Delhi, with approval from the AIIMS Institute Ethics Committee (IEC) with Ref. No. IEC-371/06.05.2022, RP-63/2022. In addition to Geriatric Medicine, the departments of Obstetrics and Gynecology, Pediatrics, Biophysics, Biochemistry, Psychiatry, Cardiac Biochemistry, Laboratory Medicine, and Biostatistics at AIIMS are also part of the study. This study will include 200 healthy participants from five cohorts (40 participants per cohort), *i*.*e*. newborns (Cohort A), adolescents (Cohort B: 10-19 years), middle-aged individuals (Cohort C: 20-59 years), young olds (Cohort D: 60-79 years) and the oldest old (Cohort E: Above 80 years). At least three age groups/ 3 generations from a single family will be counseled to participate in the study together. Recruiting people from the same families might reveal genetic factors associated with longer life spans in certain families.

Participants with severe medical conditions such as cancer, immediate surgery, immunomodulatory disease, and neurodegenerative disease that can significantly affect the biomarker profile will be excluded. However, participants with the most common lifestyle diseases, such as well-controlled hypertension, diabetes, and mild knee osteoarthritis, will be included in the study.

### Recruitment plan

Considering the challenge of recruiting at least three members from different generations within a single family, various strategies have been adopted to recruit study participants. First, individuals are contacted through emails, WhatsApp groups, phone calls, and pamphlets to encourage participation. To enhance public participation, a Media brief related to the project was published in the leading newspaper of the NCR region; hence, any family willing to participate can be included. Moreover, an E-banner was displayed at several metro stations to make people aware of ageing and persuade them to participate in the study. An awareness camp and discussion forum was organized for Resident Welfare Associations (RWAs) to provide a better understanding of our project and its need for society.

For Cohort A (newborns), pregnant mothers attending the Obstetrics and Gynecology department will be counseled to participate in this project. If they provide consent, cord blood samples of their neonates will be collected at the time of delivery.

### Informed consent and demographics

Participants volunteering for the study will be evaluated for eligibility. If at least three generations in the family are willing to participate, consent will be obtained, and all participants will be assigned to different cohorts based on their age. After enrolling in different cohorts, each one will get an AIIMS ID and project ID.

- **Consent** First, we provide each participant with a Participant Information Sheet (PIS) to know about the project, its conduction and related risks and benefits. These PIS are designed in such a manner that every participant can understand this irrespective of their age. Thereafter, they are made to sign a Participant Informed Consent Form (PICF) to participate in the study. For minors up to 12 years of age, their parents will sign PICF – Legally Authorized Representative (LAR). For the participants aged 13 to 18 years, PICF-LAR and an assent form will be signed by parents and the participants, respectively.
- **Demographics** The participant’s demographic data will include non-clinical data about a participant, consisting of name, sex, age, phone no., address, education, and occupation. Demographics are one of the most imperative parts of patient records as they are unique to each participant.

### Assessment Road map

Various health domains **(Table 1)** will be evaluated to estimate biological age.

**Table 1:** List of domains for ageing study.

|  | **Study Domains** |
| --- | --- |
| Patients' details and lifestyle habits | • Socio-Economic Status<br>• International Physical Activity Questionnaire (IPAQ)<br>• Social well-being scale<br>• Family satisfaction Scale<br>• Substance abuse scale<br>• Nutritional behaviour<br>• Sleep assessment<br>• Spiritual well being<br>• Perceived self-health<br>• Attitude towards ageing |
| Cognitive Assessments | • Digit span test<br>• The Stroop Color and Word test<br>• The Wisconsin Card Sorting Test<br>• Rey Auditory Verbal Learning Test |
| Clinical health indicators | • Present and past medical history<br>• Morbidities and comorbidities<br>• Current medication<br>• BP and pulse rate<br>• Blood biochemistry |
| Psychological Distress | • Depression Anxiety Stress Scales- Short Form (DASS-21) |
| Physiological/physical function | • Six-minute walk test<br>• VO2 max<br>• Handgrip strength test<br>• Pegboard test<br>• Short Physical Performance Battery<br>• Bioelectrical Impedance Analysis |
| Molecular biomarkers | • Soluble blood proteins<br>• Telomere attrition<br>• DNA methylation |
| Microbiomics | • Gut microbiome metagenome |
| Personalized real-time wearable data | • Physical activity monitoring<br>• Sleep patterns<br>• Stress index<br>• Heart rate variation<br>• Gait speed. |

### Patients’ details and Lifestyle habits

- **Socio-Economic Status (SES):** The Kuppuswamy Scale is a composite score of the education and occupation of the head of the family along with the family’s monthly income, which yields a score of 3-29 (16). It will assess and classify the study populations into high, middle, and low socio-economic status.
- **Physical health and Lifestyle assessments:** Each participant’s physical activity, social well-being, family support, and substance use will be documented to evaluate their approach toward healthy life. The International Physical Activity Questionnaire (IPAQ) will assess all activities that keep them physically fit (17). Social well-being will be measured using a 7-point frequency scale starting from never, alternate days, twice a week, weekly, fortnightly, monthly, and daily. We will use the Family Satisfaction Scale (only for Cohort C, D, and E) to know how confident the participant feels about their family bond (18). Substance abuse, if any, will be recorded through the brief screener of alcohol, tobacco, and other drugs (19).
- **Nutritional Behavior:** Nutritional Behavior will be recorded through the 24-hour Diet Recall Questionnaire (20). Nutritional assessment will be done through the online application software DietCal-A tool for dietary assessment and planning.
- **Sleep assessments:** Sleep is undoubtedly the most vital aspect of overall health. To evaluate participants’ sleep quality, we will administer the Global Sleep Assessment Questionnaire (GSAQ) (21).
- **Spiritual Well-Being:** India being a multi-religious and multi-ethnic country, it is intricate to assess all ideologies through a questionnaire. Therefore, Spiritual inclination will be measured using a self-perceived 10-point numerical rating scale.
- **Perceived Self-health:** Participants’ Perceptions of their health will be evaluated on the scale of poor, fair, good, very good, and excellent.
- **Perception towards ageing and related stress:** The study will quantitatively evaluate (for cohorts C And D) people’s perception towards ageing through the Attitude to Ageing Questionnaire-Short Form (AAQ-SF) (22).

### Cognitive assessments

- Cognitive ageing is the age-related changes in the cognitive function typically occurring when people age. However, comparing executive cognitive function in various age groups is imperative to see the effect of cognitive decline with age.
- A digit span test will measure Working Memory (23).
- The Stroop Color and Word test will be used to assess selective attention and the ability to control inhibition (24).
- Rey Auditory Verbal Learning Test will be administered to assess episodic memory (25).
- The Wisconsin Card Sorting Test will be taken to measure abstract thinking and executive functioning on every participant (26).

### Clinical health indicators

- **Medical History:** The clinical history of all participants will be recorded. This database will help us to correlate biological age with the present and past clinical profiles of participants. We primarily focus on vital signs, significant morbidities, co morbidities, disease history, and present or past medication.
- **BP and pulse rate:** Systolic and diastolic blood pressure and pulse rate will be measured using a brachial pressure cuff for all participants.
- **Blood Biochemistry:** To understand bodily function, internal balance, and overall health, we will do routine blood tests of each participant. It will include a Complete Blood Count (CBC), Liver Function Test (LFT), Renal Function Test (RFT), Lipid Profile, Thyroid Profile, vitamin D, inflammation by C - C-reactive protein (CRP), and Insulin. Peripheral blood (up to 20 mL) will be drawn into Vacutainer plain tubes and EDTA tubes from the antecubital vein by a certified phlebotomist, with the participant having fasted overnight. These samples are analysed immediately according to well-established protocols for haematology, coagulation, and biochemistry tests.

### Psychological distress

- Participants will be assessed based on their emotional health using Depression Anxiety Stress Scales-Short Form (DASS-21) (27).

### Physiological/physical

- We would assess the participants of various age groups with different tests to observe the age-related changes in their functional competence.
- Six Minute Walk Test (6MWT) will be done to assess exercise capacity (28).
- Maximal oxygen uptake indicating physical fitness will be evaluated by measuring VO2 max through the Cardiopulmonary Exercise Test (CPET) (29).
- Muscular strength will be checked by the Handgrip Strength Test using Jamer’s Hydraulic Hand dynamometer (30).
- A Purdue pegboard test will assess motor coordination in both hands. It is a test that measures finger dexterity, gross movements, and coordination of both hands (31).
- A Short Physical Performance Battery (SPPB) will be used to check the participants’ balance, gait, strength, and endurance (32).
- Bioelectrical impedance analysis (BIA) will estimate body fat and muscle mass. Tanita MC-980MA Plus body composition analyses will be used for the assessment (33).

### Molecular biomarkers

- **Soluble blood proteins:** The blood samples collected from the study subjects will be analyzed for the quantification of BubR1, SIRT1, HSF1, Akt, mTORC1, FOXO3A, AMPK, PGC1α, NFκB and P16INK4a proteins in serum by Surface Plasmon Resonance (SPR) technology (34). **(Table 2)**
- In brief, peripheral blood will be collected in BD Vacutainer® Plus Plastic Serum Tubes, which will be kept undisturbed at room temperature for 15-30 minutes to allow blood clotting. Blood will then be centrifuged at 1000–2000 x g for 10 minutes in a refrigerated centrifuge (Eppendorf, Germany) to obtain serum as suspension. The serum will be immediately transferred into a clean polypropylene tube in aliquots of 0.5 ml and stored at –80°C for future SPR analysis.

**Table 2:** Ageing biomarkers respective to ageing hallmarks.

| S. No. | Hallmarks of Ageing | Respective Biomarker of Ageing | Reason for Selection |
| --- | --- | --- | --- |
| 1. | Genomic instability | Mitotic checkpoint<br>serine/threonine-protein kinase<br>BUB1 beta (BubR1) (35) | • Participates in cell cycle regulation<br>• Decreases with age |
| 2. | Telomere attrition | Telomere Attrition assay (36) | • Length of telomeres decreases with age |
| 3. | Epigenetic alterations | NAD-dependent protein<br>deacetylase sirtuin-1(SIRT1)<br>(37) | • Participates in DNA repair<br>• Decreases with age |
| 4. | Loss of proteostasis | Heat Shock Factor 1 (HSF1)<br>(38) | • Protects from Proteotoxic stress<br>• Decreases with age |
| 5. | Deregulated nutrient-sensing | • RAC-alpha serine/threonine-protein kinase (Akt) (39)<br>• Mechanistic target of rapamycin complex 1 (mTORC1) (40) | • Maintains metabolic homeostasis<br>• Increases with age, thereby affecting nutrient metabolism. |
| 6. | Mitochondrial dysfunction | • Peroxisome proliferator-activated receptor gamma coactivator 1-alpha (PGC-1α) (41)<br>• Forkhead box protein O3 (FOXO3a) (42) | • Participates in mitochondrial biogenesis and energy homeostasis<br>• Decreases with age |
| 7. | Cellular senescence | • Cyclin-Dependent Kinase Inhibitor 2A (p16INK4a) (43) | • Inhibits the cell cycle by irreversible cell cycle arrest<br>• Increases with age |
| 9. | Altered intercellular communication | • IL-6 (44)<br>• IL-13 | • Markers of Inflammageing<br>• Increases with age |
| 10. | Chronic inflammation | Nuclear factor NF-kappa-B p50 subunit (NF kappa b) (45) | • Activates in the presence of cellular damage and stress<br>• Increases with age leading to long-term inflammation |
| 11. | Disabled macroautophagy | 5'AMP-activated protein kinase catalytic subunit alpha-1 (AMPKα) (46) | • Initiates autophagy and regulates lysosomal biogenesis<br>• Decreases with age |

Quantification by SPR will be performed using the BIAcore-3000 [Pharmacia Biosensor AB, Uppsala, Sweden], a biosensor-based system for real-time monitoring of specific interaction analysis of target molecules. Primary antibodies will be immobilized on the flow cell of the CM5 sensor chip via an amine coupling kit (Pharmacia Biosensor AB). Standard graphs will be prepared by passing different known concentrations of pure re-recombinant proteins over respective flow cells of the sensor chip and respective response unit (RU) values obtained. The serum will be passed over the immobilized antibodies. The RU for each sample was recorded, and the serum concentration in different subjects was derived from the standard curve.

- **Telomere attrition:** The shortening of telomeres with age will be evaluated through quantitative polymerase chain reaction (QPCR) on genomic DNA. Leukocyte telomere length will be measured usingthe monochrome multiplex quantitative PCR (MMqPCR) method described by Cawthon (47).

Peripheral blood will be collected in BD Vacutainer® Plus EDTA Tubes. PBMCs will be isolated from the peripheral blood of study subjects by the Ficoll Hypaque gradient centrifugation method (GE Healthcare, Sweden). The PBMC pellet will be immediately transferred into a clean polypropylene tube and stored at –80°C for future DNA extraction. At the time of assay, genomic DNA will be extracted from the frozen PBMCs of individual participants with the QIAamp® DNA Mini Kit (QIAGEN Singapore Pte. Ltd.).

DNA extracted from blood samples will be taken in 1×master mix consisting of 0.5×SYBR Green I (Invitrogen, CA, USA); 0.5AmpliTaq Gold DNA polymerase (Applied Biosystems, CA, USA), 10 mM Tris-HCl, 50 mM KCl, three mM MgCl2 (Invitrogen), 0.2 mM each dNTP (MBI Fermentas, Hanover, MD, USA), one mM DTT (Sigma, Germany), 1 M betaine (Sigma, Germany) and nuclease-free water in a final volume of 25μl and added into reaction wells of 96-well PCR plate (Axygen, CA, USA) compatible with Real-Time PCR detection system (IQ5, software version 2.0, Bio-Rad Laboratories Inc., CA, USA) and amplified. Each run will amplify one non-template control and two positive controls in duplicates. For the multiplex qPCR, HPLC-purified forward and reverse telomere primer pair (final concentration of 300 nM each) and single-copy gene, albumin primer pair (final concentration 300 nM each) will be included in each reaction. Thirty-five cycles of PCR will be carried out, and the signal will be acquired using Bio-Rad iQ5 software version 2.0.T. After the run is completed, the Ct values of telomere and albumin gene will be transferred to the spreadsheet separately to generate standard curves from which the copy number for telomere and single-copy gene values will be generated. The ratio of the copy number of telomeres and the single-copy gene will be determined, indicative of the average telomere length per cell.

- **DNA Methylation:** Peripheral blood will be collected in BD Vacutainer® Plus EDTA Tubes. PBMCs will be isolated from the peripheral blood of study subjects by the Ficoll Hypaque gradient centrifugation method (GE Healthcare, Sweden). The PBMC pellet will be immediately transferred into a clean polypropylene tube and stored at – 80°C for future DNA extraction. At the time of assay, DNA will be extracted from the frozen PBMCs of individual participants with the QIAamp® DNA Mini Kit (QIAGEN Singapore Pte. Ltd.). After a quality check, all DNA samples will measure the methylation pattern. The quality of the genomic DNA will be assessed through A260/A280 and A260/A230 spectrophotometry ratios and the concentrations of DNA isolated. The Illumina Infinium Methylation EPIC v2.0 Bead Chip Array (Illumina, Inc. San Diego, CA, USA) will be used for high throughput genome-wide DNA methylation measurement. The 935 K enables reliable and reproducible evaluation of over 935,000 CpG methylation sites, offering extended genomic coverage of gene regulatory regions.

### Microbiomics

Stool samples for gut microbiota analysis are collected for cutting-edge metagenomic sequencing to get personalized insights into the gut’s ecosystem, microbial richness, and diversity. Microbiome metagenome analyses will includehigh-throughput DNA/RNA sequencing methodologies (e.g. Illumina Mi/HiSeq) for sequencing, data analysis, specific protocols, and bioinformatics pipelines.

Participants will be provided with an empty stool collection container during the screening visit and return the stool container during the recruitment visit to be stored at –80°C for future analysis.

### Wearable devices

Real-time health monitoring of study participants will be done through wearable devices. The device would monitor the study participants’ physical activity; sleep patterns, stress index, heart rate variation, and gait speed and generate real-time data.

### Data analysis, integration, and modelling

- **Bioinformatics and multi-model data integration: Bioinformatics and multi-model data integration:** Considering the multifaceted nature of ageing, a single data modality may be insufficient to capture the complexity of biological ageing. Data integration is an essential and indispensable field of ageing research. We will integrate data, including epigenetics, phenomics, clinical tests, blood biomarkers, and telomere data, to develop an integrative biomarker that can better represent biological ageing, improve predictability and more accurately reflect the pace of biological ageing. Most importantly, handling this large set of big data, which can be continuous and discrete, requires the modern analytics of machine learning. Depending on the nature of the data, we will deploy machine learning methods and, in particular, deep learning approaches to integrate various modalities. The idea is to develop a predictive algorithm for biological ageing.
- **Assessment of the pace of BA:** DunedinPACE is the most advanced third-generation epigenetic clock that has recently been described to quantify the pace of biological ageing (12,48). After developing a predictive model for BA, the next step would be the assessment of the pace of BA. After 5 years, we would assess all parameters again for all participants to measure the pace of ageing as per the DunedinPACE protocol.
- **Statistical Analysis:** Data will be recorded on a pre-designed performance and managed digitally with the Research Electronic Data Capture (REDCap) data collection tool (49). For nutritional assessment, we are using the online application software DietCal. Study subjects’ demographic, clinical, physiological, and blood-based data will be compared among the cohorts. Quantitative variables will be summarized by mean [±standard deviation (SD)]/ median [interquartile range (IQR)], as appropriate. One-way ANOVA/ Kruskal Wallis test will compare mean/ median/ among the cohorts. Post hoc ANOVA will be used if required. Categorical variables will be summarized by frequency (percentage). Chi-square test/ Test of proportions will be used to compare frequencies between the groups. Fisher’s exact test will investigate associations between two categorical or binary variables, if any. Stata 18.0 and GraphPad Prism 10 statistical software will be used for data analysis and graphical representation. The association between different biomarkers will be investigated both within and between cohorts. In this study, p-value ≤ 0.05 will be considered statistically significant.

Schematic representation of complete protocol is given in **Figure 1**.

**Figure 1.**
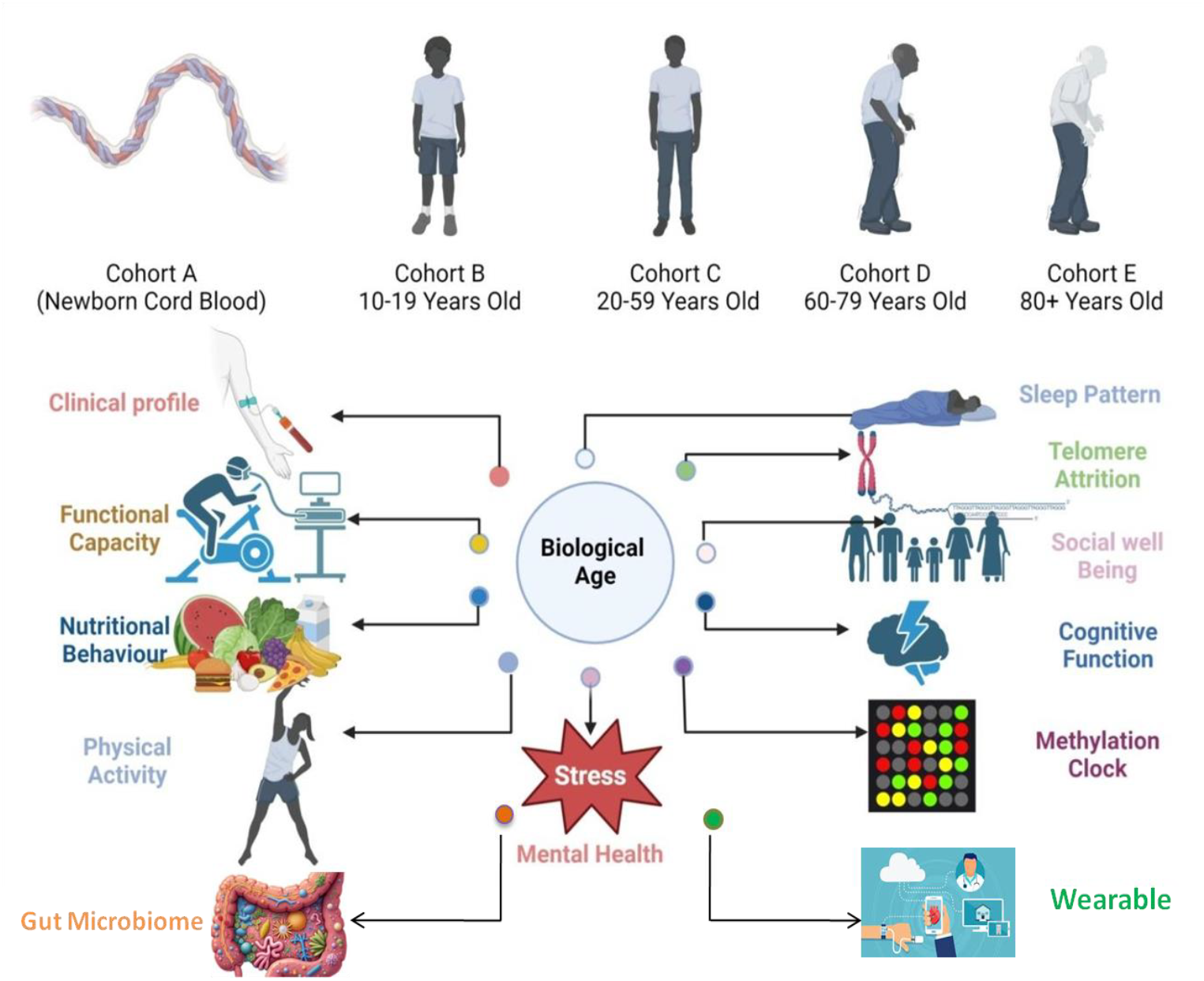
Schematic Diagram for Study Design

## Discussion

Understanding the biological determinants of healthy ageing is a critical step in maintaining the longevity of the elderly population. This study protocol outlines a comprehensive, multi-cohort, across-lifespan approach to identify and evaluate ageing biomarkers among diverse populations in the Delhi NCR region. This will be one of the first of this kind of study globally to look for multi-parametric variables representing genetic, epigenetic, metabolic, cognitive, physical, psychological, and inflammatory ageing processes across the five generations, covering all the hallmarks of ageing. Moreover, the integration of various biomarkers to generate a multi-model algorithm of biological age will provide an understanding of the ageing system as a whole rather than focusing on individual aspects. Thus, this study is expected to provide a more nuanced understanding of ageing and longevity within the unique socio-cultural context of India.

Characterizing specific biomarkers of ageing measures is vital, offering great potential for enabling human longevity intervention trials and personalized clinical decision-making. As per the FDA-BEST (The Biomarkers, EndpointS and other Tools) glossary, ageing biomarkers can broadly be classified as molecular, biological, functional, clinical, physiological, histologic, radiographic, phenotypic, and digital markers of ageing (50). This study is designed to encompass most, if not all, of these biomarkers.

This study has several strengths. 1) It has a heterogeneous sample population, as Delhi has significant numbers of immigrants from all across India. 2) Age-sensitive heterogenous biomarkers covering all aspects of ageing. 3) 5-year follow-up explaining the ageing pace by measuring the damage rate over the ageing years. 4) Wearable devices tracking the health metrics of participants in real-time, leading to a “personalized ageing” approach.

The study may face the following challenges as well: Firstly, bringing healthy individuals from the community-dwelling to the hospital for detailed analysis will be difficult as AIIMS, New Delhi, is an extremely busy tertiary care hospital. This study will follow the subjects with similar biomarker assessments once at baseline and after 5 years. It will be difficult to predict life span or mortality with this small follow-up, even though a complex AI-based algorithm would help our analysis. There will be chances of attrition during follow-up, and handling the wearable devices for technology-naive people may be difficult.

Future studies involving various omics data (transcriptomics, proteomics, and metabolomics) generated on specific cells or tissue may prove valuable. Cell-type- and tissue-specific biomarkers could elucidate organ-specific ageing, reflecting the idea of “Ageotyping”. Moreover, long-term follow-ups with larger sample sizes might complement the outcome of our research.

## Conclusion

This study will present a comparative assessment of age-related biomarkers among the cohorts of varied age groups and the role of epigenetic changes, especially methylation and telomere attrition, in cellular ageing. The Role of the genetic makeup of a family in ageing will also be discussed. Identification of biological ageing markers and health status will lead to more robust evidence to facilitate the development and targeting of interventions to improve health and avert high-cost dependency. Still, it will also aid investigations of the links between ageing and disease. We aim to develop an affordable, feasible, and reproducible integrative biomarker panel that can manifest health outcomes of the ageing process. This study also has potential translational implications. Identifying longevity biomarkers could pave the way for targeted strategies to promote healthy ageing.

## Data Availability

All data produced in the present study are available upon reasonable request to the authors

## Acknowledgements

The authors would like to acknowledge Dr. Saad Mustafa, Ms. Mukti Gupta, for their intellectual input in protocol development.

## Author contribution

Conceptualization: Prasun Chatterjee; Methodology: Prasun Chatterjee, Rashi Jain; Lata Rani; Writing—original draft preparation: Prasun Chatterjee, Rashi Jain, Pooja; Writing— review and editing: Prasun Chatterjee, Rashi Jain, Pooja, Avinash Chakrawarty, Lata Rani, Sharmistha Dey, Rashmita Pradhan, Vidushi Kulshrestha, Lakshmy Ramakrishnan, Prasun Chatterjee; Supervision: Prasun Chatterjee.

## Data availability

Since this is a protocol, no data generation or analyses have been performed in this article. For further inquiries, please contact the corresponding author.

## References

1. da Costa JP, Vitorino R, Silva GM, Vogel C, Duarte AC, Rocha-Santos T. A synopsis on aging-Theories, mechanisms and future prospects. Ageing Res Rev. 2016 Aug;29:90–112.

2. Zhang Q. An interpretable biological age. Lancet Healthy Longev. 2023 Dec 1;4(12):e662–3.

3. Bortz J, Guariglia A, Klaric L, Tang D, Ward P, Geer M, et al. Biological age estimation using circulating blood biomarkers. Commun Biol. 2023 Oct 26;6(1):1089.

4. American Federation for Aging Research [Internet]. [cited 2025 Feb 11]. FAST Initiative. Available from: https://www.afar.org/fast-initiative

5. Galkin F, Mamoshina P, Aliper A, de Magalhães JP, Gladyshev VN, Zhavoronkov A. Biohorology and biomarkers of aging: Current state-of-the-art, challenges and opportunities. Ageing Res Rev. 2020 Jul 1;60:101050.

6. Aanes H, Bleka Ø, Dahlberg PS, Carm KT, Lehtimäki T, Raitakari O, et al. A new blood based epigenetic age predictor for adolescents and young adults. Sci Rep. 2023 Feb 9;13(1):2303.

7. Chen BH, Marioni RE, Colicino E, Peters MJ, Ward-Caviness CK, Tsai PC, et al. DNA methylation-based measures of biological age: meta-analysis predicting time to death. Aging. 2016 Sep 28;8(9):1844–65.

8. Horvath S. DNA methylation age of human tissues and cell types. Genome Biol. 2013 Dec 10;14(10):3156.

9. Hannum G, Guinney J, Zhao L, Zhang L, Hughes G, Sadda S, et al. Genome-wide methylation profiles reveal quantitative views of human aging rates. Mol Cell. 2013 Jan 24;49(2):359–67.

10. Lu AT, Quach A, Wilson JG, Reiner AP, Aviv A, Raj K, et al. DNA methylation GrimAge strongly predicts lifespan and healthspan. Aging. 2019 Jan 21;11(2):303–27.

11. Levine ME, Lu AT, Quach A, Chen BH, Assimes TL, Bandinelli S, et al. An epigenetic biomarker of aging for lifespan and healthspan. Aging. 2018 Apr 17;10(4):573– 91.

12. Belsky DW, Caspi A, Corcoran DL, Sugden K, Poulton R, Arseneault L, et al. DunedinPACE, a DNA methylation biomarker of the pace of aging. eLife. 11:e73420.

13. López-Otín C, Blasco MA, Partridge L, Serrano M, Kroemer G. The Hallmarks of Aging. Cell. 2013 Jun 6;153(6):1194–217.

14. López-Otín C, Blasco MA, Partridge L, Serrano M, Kroemer G. Hallmarks of aging: An expanding universe. Cell. 2023 Jan 19;186(2):243–78.

15. Wang J, Chen C, Zhou J, Ye L, Li Y, Xu L, et al. Healthy lifestyle in late-life, longevity genes, and life expectancy among older adults: a 20-year, population-based, prospective cohort study. Lancet Healthy Longev. 2023 Oct 1;4(10):e535–43.

16. Radhakrishnan M, Nagaraja SB. Modified Kuppuswamy socioeconomic scale 2023: stratification and updates. Int J Community Med Public Health. 2023 Oct 31;10(11):4415– 8.

17. Lee PH, Macfarlane DJ, Lam T, Stewart SM. Validity of the international physical activity questionnaire short form (IPAQ-SF): A systematic review. Int J Behav Nutr Phys Act. 2011 Oct 21;8(1):115.

18. Zabriskie RB, Ward PJ. Satisfaction With Family Life Scale. Marriage Fam Rev. 2013 Jul;49(5):446–63.

19. Levy S, Brogna M, Minegishi M, Subramaniam G, McCormack J, Kline M, et al. Assessment of Screening Tools to Identify Substance Use Disorders Among Adolescents. JAMA Netw Open. 2023 May 22;6(5):e2314422.

20. j4-hour Dietary Recall (24HR) At a Glance | Dietary Assessment Primer [Internet]. [cited 2025 Feb 12]. Available from: https://dietassessmentprimer.cancer.gov/profiles/recall/

21. Roth T, Zammit G, Kushida C, Doghramji K, Mathias SD, Wong JM, et al. A new questionnaire to detect sleep disorders. Sleep Med. 2002 Mar;3(2):99–108.

22. Laidlaw K, Kishita N, Shenkin SD, Power MJ. Development of a short form of the Attitudes to Ageing Questionnaire (AAQ). Int J Geriatr Psychiatry. 2018 Jan;33(1):113– 21.

23. Mefferd Jr. RB, Wieland BA, James WE. Repetitive psychometric measures: Digit span. Psychol Rep. 1966;18(1):3–10.

24. Stroop JR. Studies of interference in serial verbal reactions. J Exp Psychol. 1935;18(6):643–62.

25. Schmidt M. Rey Auditory Verbal Learning Test: RAVLT : a Handbook. Western Psychological Services; 1996. 137 p.

26. Grant DA, Berg EA. Wisconsin Card Sorting Test [Internet]. 2014 [cited 2024 Jul 24]. Available from: https://doi.apa.org/doi/10.1037/t31298-000

27. Lovibond SH, Lovibond PF. Depression Anxiety Stress Scales [Internet]. 2011 [cited 2024 Jul 24]. Available from: https://doi.apa.org/doi/10.1037/t01004-000

28. Casanova C, Celli BR, Barria P, Casas A, Cote C, Torres JP de, et al. The 6-min walk distance in healthy subjects: reference standards from seven countries. Eur Respir J. 2011 Jan 1;37(1):150–6.

29. McGreevy KM, Radak Z, Torma F, Jokai M, Lu AT, Belsky DW, et al. DNAmFitAge: biological age indicator incorporating physical fitness. Aging. 2023 Feb 22;15(10):3904–38.

30. Bohannon RW. Grip Strength: An Indispensable Biomarker For Older Adults. Clin Interv Aging. 2019 Oct 1;14:1681–91.

31. Purdue Pegboard Test (PPT) – Strokengine [Internet]. [cited 2025 Feb 12]. Available from: https://strokengine.ca/en/assessments/purdue-pegboard-test-ppt/

32. Guralnik JM, Simonsick EM, Ferrucci L, Glynn RJ, Berkman LF, Blazer DG, et al. A short physical performance battery assessing lower extremity function: association with self-reported disability and prediction of mortality and nursing home admission. J Gerontol. 1994 Mar;49(2):M85–94.

33. Mandalà C, Veronese N, Dominguez LJ, Candore G, Accardi G, Smith L, et al. Use of bioelectrical impedance analysis in centenarians: a systematic review. Aging Clin Exp Res. 2023;35(1):1–7.

34. Nguyen HH, Park J, Kang S, Kim M. Surface Plasmon Resonance: A Versatile Technique for Biosensor Applications. Sensors. 2015 May 5;15(5):10481–510.

35. Baker DJ, Dawlaty MM, Wijshake T, Jeganathan KB, Malureanu L, van Ree JH, et al. Increased expression of BubR1 protects against aneuploidy and cancer and extends healthy lifespan. Nat Cell Biol. 2013 Jan;15(1):96–102.

36. Vaiserman A, Krasnienkov D. Telomere Length as a Marker of Biological Age: State-of-the-Art, Open Issues, and Future Perspectives. Front Genet [Internet]. 2021 Jan 21 [cited 2024 Jul 26];11. Available from: https://www.frontiersin.org/journals/genetics/articles/10.3389/fgene.2020.630186/full

37. Rogina B, Tissenbaum HA. SIRT1, resveratrol and aging. Front Genet [Internet]. 2024 May 9 [cited 2025 Feb 12];15. Available from: https://www.frontiersin.org/journals/genetics/articles/10.3389/fgene.2024.1393181/full

38. Lazaro-Pena MI, Ward ZC, Yang S, Strohm A, Merrill AK, Soto CA, et al. HSF-1: Guardian of the Proteome Through Integration of Longevity Signals to the Proteostatic Network. Front Aging [Internet]. 2022 Jul 8 [cited 2025 Feb 12];3. Available from: https://www.frontiersin.org/journals/aging/articles/10.3389/fragi.2022.861686/full/www.frontiersin.org/journals/aging/articles/10.3389/fragi.2022.861686/full

39. Chen YR, Li YH, Hsieh TC, Wang CM, Cheng KC, Wang L, et al. Aging-induced Akt activation involves in aging-related pathologies and Aβ-induced toxicity. Aging Cell. 2019 Aug;18(4):e12989.

40. Ortega-Molina A, Lebrero-Fernández C, Sanz A, Calvo-Rubio M, Deleyto-Seldas N, de Prado-Rivas L, et al. A mild increase in nutrient signaling to mTORC1 in mice leads to parenchymal damage, myeloid inflammation and shortened lifespan. Nat Aging. 2024 Aug;4(8):1102–20.

41. Wenz T. Mitochondria and PGC-1α in Aging and Age-Associated Diseases. J Aging Res. 2011 May 5;2011:810619.

42. Zhao Y, Liu YS. Longevity Factor FOXO3: A Key Regulator in Aging-Related Vascular Diseases. Front Cardiovasc Med. 2021 Dec 23;8:778674.

43. LaPak KM, Burd CE. The Molecular Balancing Act of p16INK4a in Cancer and Aging. Mol Cancer Res MCR. 2014 Feb;12(2):167–83.

44. Tylutka A, Walas Ł, Zembron-Lacny A. Level of IL-6, TNF, and IL-1β and age-related diseases: a systematic review and meta-analysis. Front Immunol. 2024 Mar 1;15:1330386.

45. Tilstra JS, Clauson CL, Niedernhofer LJ, Robbins PD. NF-κB in Aging and Disease. Aging Dis. 2011 Dec 2;2(6):449–65.

46. Burkewitz K, Zhang Y, Mair WB. AMPK at the Nexus of Energetics and Aging. Cell Metab. 2014 Jul 1;20(1):10–25.

47. Cawthon RM. Telomere measurement by quantitative PCR. Nucleic Acids Res. 2002 May 15;30(10):e47.

48. Salameh Y, Bejaoui Y, El Hajj N. DNA Methylation Biomarkers in Aging and Age-Related Diseases. Front Genet [Internet]. 2020 Mar 10 [cited 2024 Jul 26];11. Available from: https://www.frontiersin.org/journals/genetics/articles/10.3389/fgene.2020.00171/full

49. Harris PA, Taylor R, Thielke R, Payne J, Gonzalez N, Conde JG. Research electronic data capture (REDCap)--a metadata-driven methodology and workflow process for providing translational research informatics support. J Biomed Inform. 2009 Apr;42(2):377–81.

50. FDA-NIH Biomarker Working Group. BEST (Biomarkers, EndpointS, and other Tools) Resource [Internet]. Silver Spring (MD): Food and Drug Administration (US); 2016 [cited 2025 Feb 12]. Available from: http://www.ncbi.nlm.nih.gov/books/NBK326791/

